# Malaria Infection in Patients with Sickle Cell Disease in Nigeria: Association with Markers of Hyposplenism

**DOI:** 10.1101/2023.05.08.23289666

**Authors:** Adama I Ladu, Mairo Y Kadaura, Mohammed Dauda, Abubakar Sadiq Baba, Nasir Garba Zango, Caroline Jeffery, Abubakar Farate, Adekunle Adekile, Imelda Bates

## Abstract

**Background:** Malaria is considered an important cause of morbidity and mortality among people living with sickle cell disease (SCD). This has partly been attributed to the loss of splenic function that occurs early in the disease process. We aimed to study the prevalence of malaria infection among Nigerian SCD patients and explore the association with spleen size and function.

**Method:** This was a hospital-based, cross-sectional study performed at the University of Maiduguri Teaching Hospital in North-Eastern Nigeria from October 2020 to November 2021. Giemsa-stained blood smears for malaria parasites, Howell-Jolly body (HJB) red cells enumeration for spleen function evaluation and ultrasonography for spleen size assessment, were performed in acutely-ill SCD patients. Results of malaria parasitaemia and parasite density were compared with those of steady-state SCD patients and non-SCD controls.

**Results:** A total of 394 participants consisting of 119 acutely-ill SCD patients, 167 steady-state SCD controls and 108 non-SCD controls were studied. The prevalence of *P. falciparum* parasitaemia was 51.3% in acutely-ill SCD patients, 31.7% in steady-state SCD controls and 13.0% in the non-SCD controls. In the SCD group, the mean parasite density was significantly higher among the acutely-ill SCD patients than the steady-state SCD controls (29,747 vs 18,563 parasites / ul; *P = 0*.*001*). Although parasitaemia prevalence was lower among the non-SCD controls, parasite density was significantly higher compared to both SCD groups (*P = 0*.*0001*). Among the acutely-ill SCD patients, the prevalence of clinical malaria and severe malaria anaemia were highest among children less than 5 years of age. Prevalence of parasitaemia (*P = 0*.*540*) and parasite density (*P = 0*.*975*) among acutely-ill SCD patients with visualized spleens on ultrasonography were not statistically different compared to those with absent spleens. Similarly, the frequency of HJB red cells among patients with parasitaemia was not significantly different compared to patients without parasitaemia (*P = 0*.*183*).

**Conclusion:** Our study highlights the frequency and role of malaria infection in acutely-ill SCD patients, especially in those younger than five years. Although we have found no evidence of an increased risk of malaria parasitaemia or parasite density with markers of hyposplenism, the role played by an underlying immunity to malaria among SCD patients is not clear. Further studies are required to elucidate the role of hyposplenism and malaria in SCD patients in malaria-endemic regions.

## Introduction

More than 300,000 infants are born worldwide with sickle cell disease (SCD) annually, and about two-thirds of these are in Sub-Saharan Africa (SSA) where malaria infection is also endemic (1). Malaria has widely been considered a major cause of morbidity and mortality among SCD patients in SSA (2-5). The heterozygous form of the sickle gene (HbAS) confers both innate and acquired protection against malaria infection (6, 7). HbAS red cells sickle preferentially when they are infected with *P. falciparum*, the infectious agent causing malaria; the parasitized sickled cells are phagocytosed by macrophages of the reticulo-endothelial system in the spleen and other organs (8). The clinically-relevant consequence of this process is that the parasite level is relatively low in Hb AS heterozygotes. It has been suggested that the degree of protection against malaria infection might be correlated with the intracellular concentration of HbS, thus the protection should be greater among the homozygotes (9). However, clinical studies of malaria among SCD patients have shown differing outcomes. A lower prevalence and density of malaria parasitaemia among individuals with HbSS compared to normal controls was reported in studies from Kenya (HbSS) (10-12), Tanzania (13) and Nigeria (14, 15), suggesting that SCD patients experience some degree of resistance to the infection than heterozygotes. Other studies have shown that homozygotes (HbSS) can develop severe and fatal complications following infection with *P. falciparum* (16-18); malaria infection was a common precipitating factor for both vaso-occlusive and haemolytic crisis.

The presence of splenic dysfunction which occurs early in the disease process through recurrent vaso-occlusion may make malaria more severe among SCD individuals compared to normal individuals (19). The spleen plays an important role in the control of *P. falciparum* parasite load even in the presence of pre-existing antibody-acquired immunity (20, 21). The absence of the spleen could thus affect an individual’s ability to clear parasitized red cells from the circulation; other organs such as the liver may take over the role of parasite clearance, but they are less efficient than the spleen (20). Cohort studies among SCD patients residing in the Northern hemisphere show that loss of splenic function occurs as early as 6 months of age and a majority of children would have reduced or absent spleen function by the age of two years (22, 23). There are few data on splenic function in the African context (24, 25), thus little is known about the increased risk of malaria associated with hyposplenism among our SCD patients (26).

While SCD patients are at an increased risk of malaria infection from hyposplenism, individuals resident in endemic regions often develop immunity following repeated episodes of infection (27, 28). A large number of the general population in malaria-endemic regions carry malaria parasites throughout life but are asymptomatic (29), however, the relevance of asymptomatic parasite carriage among SCD patients is not very clear (30, 31). The aim of our study was to determine the frequency of malaria parasitaemia and parasite density among acutely-ill SCD patients and compare it with those of steady-state SCD controls and non-SCD controls from the same environment to determine the role malaria plays in crisis. The relationships between parasitaemia prevalence and parasite density with spleen size and function among the acutely-ill patients were also evaluated to ascertain if hyposplenism is associated with increased risk of malaria.

## Methods

### Study design, setting and period

This was a hospital-based, cross-sectional study conducted at the University of Maiduguri Teaching Hospital, North-Eastern Nigeria from October 2020 to November 2021. It is a tertiary facility of 500-bed capacity, which serves as a referral hospital to Maiduguri and the surrounding township. Maiduguri is the largest city in North-East Nigeria, with an estimated population of about 800 thousand people (32). It is located in the Sahel-Savanna and is characterized by three climate patterns - a harmattan or cool-dry season (October–February), a hot season (March–May/June) and a rainy season (June or July–September) (33). Malaria is meso-endemic in Maiduguri with a seasonal transmission of 4 - 6 months during the rainy season. The prevalence of *P. falciparum*, from previous studies, ranges from 22.6% to 36% among febrile children and adults (34, 35), with the highest prevalence during the rainy seasons of August and September (74% to 84%) (34). Part of the routine care of individuals living with SCD in Nigeria is the administration of malaria chemoprophylaxis using proguanil (36).

### Study participants and data collection

All febrile and/or acutely-ill SCD patients who attended the hospital during the study period were invited to participate. The patients were divided into four age groups 1: 1 - <5 years; 2: 5 - <10 years; 3: 10 - <15 years and group 4: ≥15 years. To put our findings into context, we compared them with two control groups. The first consisted of SCD patients considered to be in steady state (37), on follow-up at the outpatient paediatric and adult haematology clinics during the study period. The second group included healthy individuals (i.e., with no SCD) consisting of medical students, children of hospital personnel and paediatric patients on post-op follow-up in the surgical clinic.

A case report form was used to obtain demographic characteristics and self-reported medical history from the patients (or their carers), including the use of insecticide-treated bed nets, anti-malaria prophylaxis, and the use of hydroxyurea in the 12 months preceding the study. Axillary temperature was measured in all patients. Malaria among the acutely-ill SCD patients was classified into three categories: (1) parasitaemia, a positive blood film (2) clinical malaria, a positive blood film in the presence of fever (defined as an axillary temperature of >37.5°C); and (3) severe malarial anaemia (SMA), haemoglobin of less than 5 g/dL in the presence of malaria parasitaemia (10, 38)

### Laboratory analysis

#### Routine blood counts

Blood was collected from all the acutely-ill SCD patients into plain tubes and ethylenediamine tetra-acetic acid (EDTA)-containing tubes. Full blood counts were performed with the use of an automated analyser (Siemens). Reticulocytes were counted using the standard method manual with methylene blue staining (39). Haemoglobin phenotyping was performed by ion-exchange high-performance liquid chromatography (HPLC) on an automatic analyser (Bio-Rad, Hercules, CA, USA). Biochemical tests were performed using a chemical analyser (Hitachi Cobas C311, Roche Instrument Centre, Rotkreuz, Switzerland).

##### Blood smears for malaria parasite analysis

Giemsa-stained thin smears for malaria parasites detection were made from EDTA blood samples not more than 4 hours after collection, air-dried and fixed in 100% methanol. Slides were then stained in 20% Giemsa (diluted in a buffer with a pH of 7.2) for 20 min. *P. falciparum* densities were assessed by counting number of asexual-stage parasites per 500 red blood cells (RBC) and expressed as parasites per microliter of whole blood. The density of parasites was calculated using each participant’s RBC count for the SCD population, or an average count of 4000 RBCs / ul of blood for the controls for whom no RBC counts were performed (40, 41).

##### Blood smears for spleen function analysis

Two blood smears for Howell-Jolly bodies (HJB) estimation were made within 4 hours of blood collection and allowed to air dry. Smears were fixed in absolute methanol for 1 minute and allowed to air dry before staining with May-Grunwald Giemsa (MGG) using the standard method (39). Four hundred RBCs were counted per smear and the HJB counts were expressed as percentages of the total red cells counted. Patients who had been transfused in the preceding 3 months were excluded from spleen function testing as this would interfere with peripheral blood film analysis.

### Ultrasonography for assessment of the spleen

All acutely-ill SCD patients were scanned using Logic P5 Premium BT11 ultrasound scanner (GE Medical Systems, USA) equipped with a low frequency (3-5MHz) curvilinear transducer as previously described (42). The scans were performed within 4 weeks of enrolment into the study. Imaging was performed by a single radiologist who assessed visualization of the spleen.

### Data processing and analysis

The data were entered into an Excel spreadsheet for cleaning and sorting and exported to Statistical Package for the Social Sciences (SPSS) (version 25; SPSS, Chicago, IL, USA) for analysis. The prevalence of malaria parasitaemia was defined as the proportion of positive blood films in all blood smears analysed in the patients or controls. Descriptive data were summarized as median or proportions, whereas between-group comparisons were analysed with logistic regression, results being presented as odds ratios (ORs) with 95% confidence intervals (95% CIs).

### Ethics statement

The study was carried out according to the Declaration of Helsinki. A signed, written informed consent was obtained from the adults and parents/guardians of the paediatric participants upon recruitment. The study was approved by the University of Maiduguri Teaching Hospital (UMTH/REC/20/606) and Liverpool School of Tropical Medicine Research (REC reference number: 20-010) Ethics Review Committee.

## Results

A total of 140 acutely-ill SCD patients (median age 13.0 years; IQR 16.0 years) were enrolled. Blood smears for malaria parasite count were available for 119 (85%); the remaining 21 samples were excluded because of poor quality. Blood smears for malaria parasite count were available for 167 steady-state SCD controls and 108 non-SCD controls. The baseline characteristics of the study population are shown in Table 1. The use of insecticide-treated bed nets (*P = 0*.*859*), malaria chemoprophylaxis (*P=0*.*350*) and hydroxyurea (*P=0*.*508*) were not significantly different between the acutely-ill SCD patients and steady-state SCD controls.

**Table 1:** Baseline characteristics of the study population.

| Parameters | Acutely-ill SCD patients | Steady-state SCD controls | Non-SCD controls |
| --- | --- | --- | --- |
| Number | 119 | 167 | 108 |
| Age (years) | 13.0 (16.0) | 11 (15.0) | 12.0 (17.8) |
| Age groups, n (%) |  |  |  |
| 1 - <5 years | 27 (22.7) | 37 (22.0) | 20 (18.5) |
| 5 - <10years | 20 (16.8) | 35 (20.8) | 22 (20.4) |
| 10 - <15 years | 27 (22.7) | 31 (21.4) | 21 (19.4) |
| ≥15 years | 45 (46.2) | 60 (35.7) | 45 (41.7) |
| Sex (male) | 55 (46.2) | 81 (48.2) | 66 (61.1) |
| Use of insecticide | 103 (86.6) | 146 (87.4) | NR |
| treated nets, n (%) |  |  |  |
| Malaria prophylaxis, n (%) | 101 (84.9) | 134 (80.2) | NR |
| Hydroxyurea therapy, n (%) | 16 (13.4) | 28 (16.8) | NR |

NR- not recorded.

### Prevalence of malaria parasitaemia and parasite density among the study participants

The prevalence of *P. falciparum* parasitaemia was 51.3% (95% Confidence interval (CI): 42% - 60%) in acutely-ill SCD patients, 31.7% (95% CI: 25% - 39%) in steady-state SCD controls and 13.0% (95% CI: 7% - 19%) in the non-SCD controls. Compared to the non-SCD control group, the odds ratio (OR) for *P. falciparum* parasitaemia was significantly higher for both the steady-state SCD controls (OR 3.12; 95% CI: 1.63 - 5.98) and acutely-ill SCD patients (7.06; 95% CI, 3.63 -13.7). Furthermore, the OR for *P. falciparum* parasitaemia was significantly higher among the acutely-ill SCD patients than the steady-state SCD controls (OR 2.26; 95% CI: 1.39 - 3.67).

Among those infected with *P. falciparum* parasites, the mean parasite density was significantly higher among the non-SCD controls (*P* = 0.0001) (Table 2). In the SCD group, the mean parasite density was significantly higher among the acutely-ill SCD patients than the steady-state SCD controls (29,747 vs 18,563 parasites/ul; *P = 0*.*001*).

**Table 2:** Comparison of parasite density across the study population.

| Variable | Geometric mean (parasite / ul) | 95% CI | P value (& post hoc analysis) |
| --- | --- | --- | --- |
| **Parasite density** |  |  |  |
| SCD patients, n=61 | 29,747 | 19,655 – 45,202 | **0.0001\*** |
| SCD-controls, n=51 | 18,563 | 13,317 – 25,769 |  |
| Non-SCD controls, n=14 | 100,512 | 50,845 – 198,693 |  |

\*Kruskal Wallis analysis; CI confidence interval; Post hoc analysis: acute-ill SCD patients vs SCD-control: 0.001; acutely-ill SCD patients vs non-SCD control: 0.0001; SCD-controls vs non-SCD controls: 0.012

### Frequency of malaria parasitaemia and parasite density across age groups among the study participants

Among the acutely-ill SCD patients, the frequency of malaria parasitaemia (*P* = 0.731) and parasite density (*P* = 0.533) was not significantly different across the age groups (Table 3). Among the steady-state SCD controls, individuals aged 15 years and above were less likely to have parasitaemia compared to those less than five years (7.5% vs 22.6%; OR 0.14; 95% CI: 0.04 - 0.49) (*P* = 0.002); however, parasite density among those with parasites detected was found to be significantly higher among the older age groups (*P = 0*.*008*). In contrast, among the non-SCD controls, the frequency of malaria parasitaemia and parasite density tended to be higher among the older individuals 10 years and above, although the difference did not reach statistical significance for either parasitaemia (*P* = 0.312) or parasite density (*P* = 0.582).

**Table 3:** Comparison of malaria parasitaemia and parasite density across age groups among the study population.

| Variables | Parasitaemia |  | Parasite density |  |
| --- | --- | --- | --- | --- |
|  | Number (%) | P values | GM (95% CI) | P values |
| **Acutely-ill SCD patients, n=61** |  |  |  |  |
| 1 - <5 years | 16 (26.2) | 0.731 | 20,737 (8449 - 50,897) | 0.533 |
| 5 - <10years | 11 (18.0) |  | 39,134 (13,285 - 115,284) |  |
| 10 - <15 years | 13 (21.3) |  | 22,862 (10,388 - 50,320) |  |
| ≥15 years | 21(34.4) |  | 39,922 (17,850 - 89,285) |  |
| **Steady-state SCD controls, n=53** |  |  |  |  |
| 1 - <5 years | 12 (22.6) | 0.0001** | 9363 (6777 - 12,936) | 0.008* |
| 5 - <10years | 24 (45.3) |  | 13,402 (9285 - 19,347) |  |
| 10 - <15 years | 13 (24.5) |  | 51, 249 (21,537 - 121,593) |  |
| ≥15 years | 4 (7.5) |  | 37,623 (4666 - 303,329) |  |
| **Non- SCD controls, n=14** |  |  |  |  |
| 1 - <5 years | 1 (7.1) | 0.312 | 96,000^^ | 0.582 |
| **5 - <10years** | 1 (7.1) |  | 192,000^^ |  |
| **10 - <15 years** | 4 (28.6) |  | 183,411 (94,128 -357,381) |  |
| **≥15 years** | 8 (57.2) |  | 69,019 (20,848 - 230,706) |  |

\*Sig.P value <0.05 - Kruskal Wallis test for parasite density across the age groups. \*\*Sig.P value <0.05 – Logistic regression analysis, 1-<5 years age group were used as the reference group; ^^CI not generated; sample size = 1. GM Geometric mean; CI Confidence interval

### Severe malaria episodes across age groups among acutely-ill SCD patients

The distribution of severe malaria episodes across age groups among the acute-ill SCD patients is shown in Table 4. Nineteen (16.0%) had clinical malaria, 15 (12.6%) had severe malaria anaemia (SMA) and seven (5.9%) had both clinical malaria and SMA. Clinical malaria was less common among older patients 15 years and above when compared to those younger than 5 years (21.1% vs 42.1%; OR 0.81; 95% CI: 0.67 - 0.98) (*P* = 0.034); however, the prevalence of SMA was not significantly different between these two age groups (33.3% vs 26.7%; OR 0.72; 95% CI: 0.18 - 2.95) (*P* = 0.646). Combined clinical malaria and SMA occurred most frequently in patients less than five-year-old (57.1%).

**Table 4:** Summary of malaria episodes across age groups among acutely-ill SCD (n-119) patients.

| Age group | Clinical malaria* | Severe malaria anaemia** | Clinical malaria and SMA |
| --- | --- | --- | --- |
| 1 - <5 years, n (%) | 8 (42.1) | 4 (26.7) | 4 (57.1) |
| 5 - <10 years, n (%) | 2 (10.5) | 2 (13.3) | 1 (14.3) |
| 10 - <15 years, n (%) | 5 (26.3) | 4 (26.7) | 2 (28.6) |
| ≥15 years, n (%) | 4 (21.1) | 5 (33.3) | 0 (0) |
| Total, n (%) | **19 (16.0)** | **15 (12.6)** | **7 (5.9)** |

\*malaria parasitaemia and temperature above 37.5° C
\*\* Malaria parasitaemia and haemoglobin level less than 5 g/dl

### Association of malaria parasitaemia with splenic parameters among acutely-ill SCD patients

Ultrasonography was available for 58 out of the 61 acutely-ill SCD patients with malaria parasitaemia. There was a tendency towards a higher prevalence of malaria parasitaemia among patients with visible spleens (n=34/58; 58.6%) (Fig. 1a) compared to those with absent spleens (n=24/58; 41.4%), however, the difference was not significant (P = 0.540). Similarly, the geometric mean parasite density in patients with visible spleens (23,100/ul) was not statistically different with those patients with absent spleens (24,300/ul) (P =0.975) (Fig.1b). The frequency of HJB red cells among patients with parasitaemia (median 1.7%, IQR 3.4) was not significantly different compared to patients without parasitaemia (median 1.5%, IQR 6.2) (*P* = 0.183).

**Fig 1.**
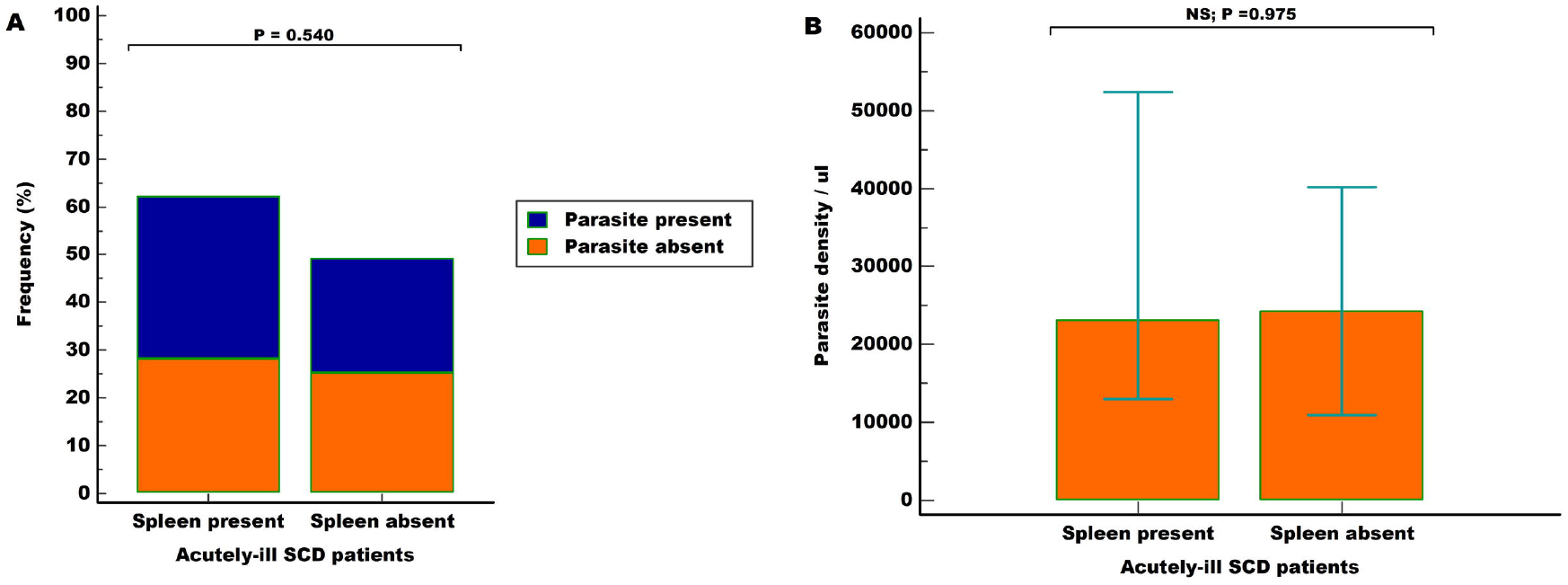
Charts showing the distribution of malaria parasitaemia and parasite density based on spleen ultrasonography among the acutely-ill SCD patients. Prevalence of malaria parasitaemia (A) (P = 0.540; Logistic regression analysis) or parasite density (B) (P=0.975; Kruskal Wallis test) was not significantly different between patients with visible or absent spleens on *ultrasonography*.

### Association of malaria parasitaemia with clinical parameters among acutely-ill SCD patients

On physical examination, pallor and jaundice were recorded in 49 (41.2%) and 42 (35.3%) patients respectively. The liver was palpable in 10 (8.4%) patients. The frequencies of a palpable liver or the presence of jaundice and pallor were not significantly different among the acutely-ill SCD patients with parasitaemia and those without (Table 5). Furthermore, the use of bed nets, antimalarial prophylaxis and hydroxyurea showed no significant difference in the prevalence of parasitaemia among the two groups. None of the laboratory parameters differed among patients with parasitaemia and those without parasitaemia.

**Table 5:**
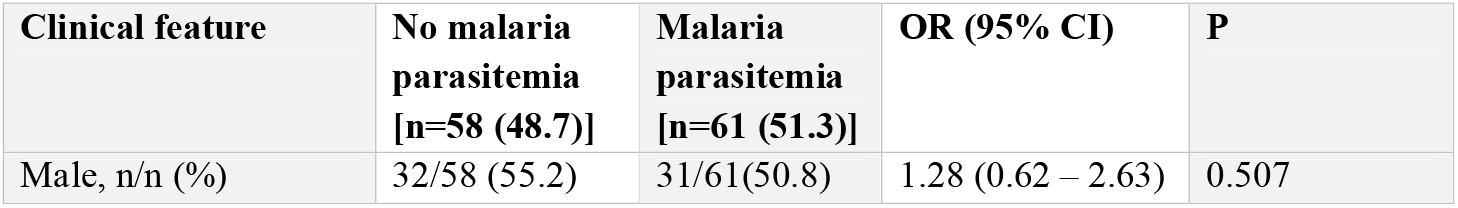
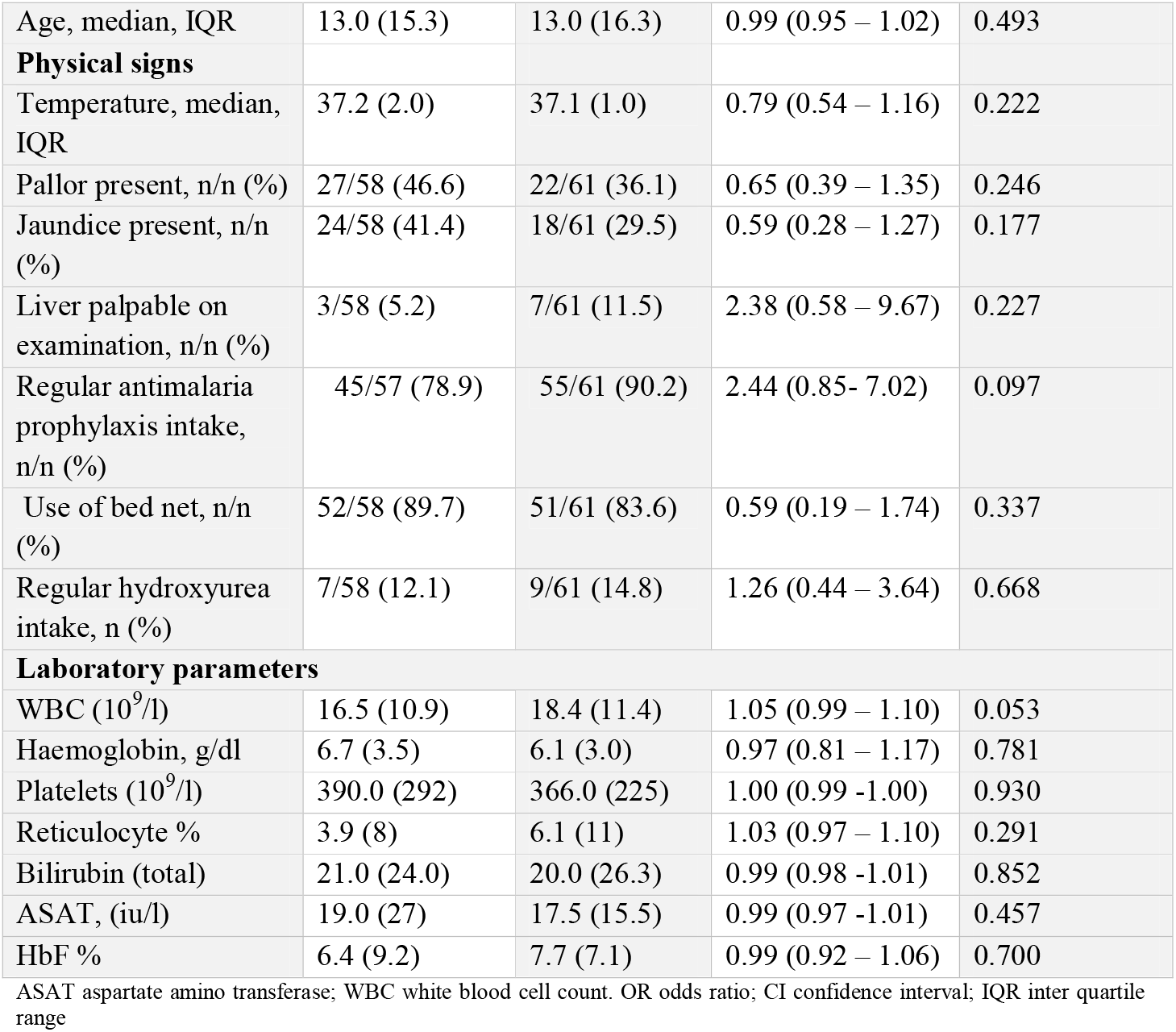
Univariate analysis for factors associated with malaria parasitaemia among acutely ill SCD patients.

## Discussion

Reports in the literature suggest that malaria is a major precipitant of crisis among SCD patients. However, since these studies were conducted in areas of high malaria transmission, where asymptomatic parasitaemia is common, and where infections from other organisms is common, the presence of parasitaemia cannot be categorically stated as the cause of the crisis (17, 18, 43, 44). The current study provided us an opportunity to evaluate the possible role of malaria among our SCD patients. We have studied malaria parasitaemia and parasite density in asymptomatic and symptomatic SCD patients and compared our results with healthy controls from the same environment. Although parasitaemia prevalence was lower among the non-SCD controls compared to both SCD groups, parasite density was significantly higher in the non-SCD controls. This may suggest a higher resistance to the parasites amongst the SCD population and maybe attributed to the high concentration of HbS in the red cells of SCD patients, which have been shown to confer protection (9, 45). However, among the SCD population, *P. falciparum* prevalence and parasites densities were higher among the acutely-ill SCD patients than steady-state SCD-controls indicating that malaria infection is an important contributor to crises. Other studies from low malaria transmission regions have shown an increased prevalence of malaria during hospitalization compared to patients in out-patient clinics (10, 38).

### Malaria burden among acutely-ill SCD patients

In malaria-endemic regions, there is a large variation in the clinical severity of *P. falciparum* infection. The malaria spectrum includes asymptomatic infection (parasitaemia), febrile illness, severe illness (including profound anaemia and cerebral complications) and death at the extreme end (46). Only a small proportion of *P. falciparum* infections progress to severe malaria; the probability of a child progressing from one event to another is as follows: asymptomatic parasitaemia (50% of each year), malaria fever twice per year; severe malaria - 3% per year, and death - 1% probability per year (46). This trend was observed in our study; we noted fewer episodes of clinical malaria (16%) compared to asymptomatic parasitaemia (51.3%). Few patients had SMA (12.5%), and fewer had a combination of clinical malaria and SMA (5.9%). This observation is similar to an earlier report from Tanzania (38). These complications were more common in the acutely-ill SCD patients less than five years of age, correlating with the high level of parasitaemia seen among the steady-state SCD controls. This finding underscores the susceptibility of under-five-year-old SCD patients to both malaria and exacerbation of their pre-existing anaemia (43), and highlights the importance of prompt and effective management of malaria among acutely-ill SCD patients to prevent progression to severe complications. Like any other infection, malaria can trigger an acute crisis in an individual with SCD. Studies have shown that both severe anaemia and death are considerably more common among SCD than non-SCD patients who were hospitalized with malaria (12, 38).

### Prevalence of parasitaemia and parasite density among steady-state SCD controls and non-SCD controls

Malaria parasitaemia was present in both our steady-state SCD controls and non-SCD controls, which may imply the presence of some degree of immunity to the parasite in the population. A high prevalence of parasitaemia was also noted among the younger (less than 10 years) steady-state SCD controls compared to the older patient population. In contrast, among the non-SCD controls, the frequency of malaria parasitaemia and parasite density showed an increasing frequency with increasing age; the prevalence was higher in patients aged 15 years and above compared to the younger patients aged five years and below.

Similarly, parasite density was higher among the older population. The small number of those less than five (n=1) and those less than 10 (n=1) years may have affected the power to detect any statistical significance among the non-SCD controls. Nevertheless, age has been shown to be an important factor that determines immunity to malaria in an endemic region (27); when a child becomes infected, he or she usually develops symptoms of the disease including fever but eventually recovers. Repeated episodes of infection among older children and adults result in the development of clinical immunity to malaria. Therefore, adults are likely to develop asymptomatic infection (27, 28, 46). As asymptomatic subjects do not seek treatment, they may serve as reservoirs of the parasites, thus contributing to the perpetual spread of the parasites (29).

### Association of malaria parasitaemia with splenic parameters among acutely-ill SCD patients

The clinical pattern and outcome of infection with *P. falciparum* may be impacted by several factors including prior exposure and immunity, intensity of transmission in the area, age, and the spleen. The spleen plays an important role in the control of *P. falciparum* parasite load (20, 21, 47). In endemic areas, fever and parasitaemia were significantly more frequent in splenectomised patients compared to subjects with intact spleen; severity and fatality of *P. falciparum* infection was increased in such patients but not to a large extent (48, 49). Mature forms of the parasite express an adhesion protein (*P. falciparum* erythrocytes membrane proteins - PfEMP1), which allows them to adhere to several cells within small vessels; by so doing, the parasites escape retention and destruction by the spleen. The PfEMP1 is a highly immunogenic molecule, and with repeated exposures, children residing in endemic areas progressively acquire antibodies against it (29). In immune patients with a spleen, as well as in splenectomised patients, antibodies to PfEMP1 prevent the sequestration of mature forms of the parasite in the microcirculation, thereby preventing severe complications. In patients with intact spleens, the mature forms are cleared by the spleen; however, in the immune individual without a spleen, the parasites are less efficiently cleared either because the microcirculation of other organs are unable to mechanically retain mature forms of the parasites or opsonisation is less efficient in the sinusoids of these organs as compared to the splenic cords (20). In our study, no significant association between the presence or absence of the spleen on ultrasonography with prevalence of parasitaemia or parasite density was noted among our acutely-ill SCD patients. Also, clinical and laboratory parameters were not different among the group with parasitaemia and those without parasitaemia. It is unclear if the presence of immunity to the parasite among our patient population accounts for this finding. The current study is the first to describe the association between malaria and splenic function (i.e. HJB) among SCD patients residing in a malaria-endemic region. The frequency of HJB red cells was not significantly different between the group with parasitaemia and those without. Splenic dysfunction has widely been reported to be associated with increased susceptibility to infections including malaria. What is not clear is whether acute malaria infection reduces spleen function as in other febrile conditions, or if it is the loss of splenic function in SCD that predisposes to the malaria infection.

### Association of malaria parasitaemia with clinical parameters among acutely-ill patients

Given the previous reports from early studies (5, 8) about the fatal consequences of malaria in SCD patients residing in malaria-endemic regions, they are routinely placed on lifelong antimalaria prophylaxis. Despite the high intake of antimalaria prophylaxis (84.9%) and use of bed nets (86.6%) among the acutely-ill SCD patients, the prevalence of malaria parasitaemia was still high. About half of the patients using bed nets (49.5%) and antimalaria prophylaxis (55%) were positive for *P. falciparum*. This is similar to the observations in several studies, where despite regular prophylaxis (with proguanil in most cases), SCD patients still developed malaria infection (17, 31, 50), calling into question the efficacy of bed nets and chemoprophylaxis. A recent Cochrane review on malaria chemoprophylaxis in SCD patients concluded it was beneficial to provide routine malaria chemoprophylaxis in SCD patients in malaria-endemic areas (51), however, decision on malaria chemoprophylaxis remains challenging (52). Other anti-malaria agents including mefloquine, amodiaquine, halofantrine and artemether-lumefantrine may be suitable for chemoprophylaxis, but they will need to be evaluated in future studies in order to provide data that can be used to develop consensus guidelines for malaria chemoprophylaxis in SCD patients (52)..

## Limitations

Our study has some limitations. Firstly, the single centre and hospital-based nature of the study conducted in an area with high malaria transmissions may affect the generalizability of our results to the general population with SCD. Secondly, we have used thin blood smears to evaluate malaria parasitaemia as opposed to thick blood films which are routinely used. The thin blood has been shown to be less sensitive at low parasitaemia (53), and may have under-estimated the prevalence of parasitaemia among our study population, especially the low rate reported among our non-SCD controls when compared to the high value reported from a previous report in the study area (34). However, an acceptable agreement between thin and thick film density measurements has been reported (54). Moreover, studies comparing parasite densities observed in thick films to those observed in thin films have reported that a large percentage of parasites can be obscured in the thick film or lost during staining and lysis of red blood cells (41, 55).

## Conclusion

In our study conducted in an area of high malaria transmission, we found that malaria parasitaemia and parasite density were more common among acutely-ill than steady-state SCD controls. We also found that severe malaria events were significantly higher among under-five than in older acutely-ill SCD patients. There was no association of malaria parasitaemia and parasite density with spleen size on ultrasonography or with splenic function.

## Data Availability

All data produced in the present study are available upon reasonable request to the authors

## Acknowledgement

The authors are grateful to the staff of Microbiology, Radiology and Haematology departments of the UMTH for their assistance during data collection, sample preparation and analysis aspect of this project. We also thank all the patients and their guardians.

## Authors’ contributions

AIL and IB conceived and designed the project. AIL, MYK, DM, NGZ and BS coordinated the patient’s recruitment sample collection and laboratory analysis. AIL and AF performed the ultrasonography. AIL performed the statistical analysis. IB, CJ and AA supervised the formal analysis. All the authors contributed significant intellectual content during the manuscript drafting or revision and accepts responsibility for the integrity of the data and accuracy of analysis.

